# A High-Coverage SARS-CoV-2 Genome Sequence Acquired by Target Capture Sequencing

**DOI:** 10.1101/2020.04.11.20061507

**Authors:** Shaoqing Wen, Chang Sun, Huanying Zheng, Ling-xiang Wang, Huan Zhang, Lirong Zou, Zhe Liu, Pan-xin Du, Lijun Liang, Xiaofang Peng, Wei Zhang, Jie Wu, Bo Lei, Changwen Ke, Fang Chen, Xiao Zhang

## Abstract

This manuscript is based on the method we developed urgently to deal with the research requirement in the conflict between achieving a complete genome sequence for the evolutionary history of SARS-CoV-2 study and the low viral RNA concentration. Here, in this manuscript, we developed a set of SARS-CoV-2 enrichment probes to increase the sensitivity of sequence-based virus detection and characterization via obtaining the comprehensive genome sequence. Following the CDC health and safety guidelines, we test the concept using the culturing supernatant contain SARS-CoV-2 particles, and its full-length sequence was used for further analysis. The fraction of SARS-CoV-2 endogenous DNA was 93.47% with Cluster Factor about 1.1, which demonstrate that the numbers of mapped reads to SARS-CoV-2 reference sequence significantly increased, compared to metagenomic sequencing technology, following SARS-CoV-2 probe enrichment. Moreover, based on the high-quality sequence, we discussed the heterozygosity and viral expression during replication of coronavirus, and its phylogenetic relationship with other selected high-quality samples from The Genome Variation Map (GVM) (on 2020/03/22). We believe this manuscript is valuable for all the researchers who are interested in using clinical warp samples to obtain the high coverage of SARS-CoV-2 genome sequence with a relatively low concentration of viral particles. This would allow the clinician to correlate the diagnostic data with molecular monitoring in viral evolutional, the most importantly, to track the functional mutation of SARS-CoV-2.

---

Ongoing outbreak of the novel coronavirus (SARS-CoV-2) disease has become a global health concern. Since a patient with pneumonia of unknown etiology were firstly reported in the city of Wuhan on 30th of Dec, 2019., Subsequently, epidemiological, clinical, radiological, laboratory and genomic findings of this virus gradually revealed by the Chinese and 00 experts around the globe [1]. At current stage, however, two topics emerged as things that needed to be addressed. Firstly, according to latest diagnostic criteria, reverse-transcriptase-polymerase-chain-reaction (RT-PCR) assays are recommended as the standard diagnosis of SARS-CoV-2-infection. Despite, present studies found that some patients had typical imaging findings, including ground-glass opacity, but negative RT-PCR testing [2]. The false-negative RT-PCR results may cause by many reasons, especially insufficient detection sensitivity in a low viral load scenario [2]. Secondly, although, by using metagenome sequencing, the complete genome sequence, taxonomic position, potential intermediate hosts, and evolutionary history of SARS-CoV-2 have been already reported [3-6], it is more important to monitor the virus mutation and its influence on disease severity and progression. Necessitating the full-length of SARS-CoV-2 genome, metagenome sequencing technology is the latest and most comprehensive approach but still costly. Moreover, in metagenome sequencing library, there are human (host) nucleic acid contamination cannot be eliminated. Inevitably, the carrier RNA introduced by commercial RNA extraction kits presented as contamination in library preparation stage, which indeed impairs the amount of viral sequence readout.

In the context, we developed a set of SARS-CoV-2 enrichment probes, by utilizing hybridization capture technology, to increase the sensitivity of sequence-based virus detection and characterization. This method was first used to enrich sequence targets from the human genome [7] and then from vertebrate virome [8]. The enrichment probe set contains 502 ssDNA biotin-labelled probes at 2X tiling designed based on all available SARS-CoV-2 viral sequences, downloaded from the GISAID (Global Initiative on Sharing All Influenza Data; https://www.gisaid.org/) on 2/1/2020, and it can be used to enrich for SARS-CoV-2 sequences without prior knowledge of type or subtype. Additionally, the probes for human housekeeping genes (GAPDH, PCBP1, EIF3L, POLR2A, EIF3A, TGOLN2, TCEB3, CDK12, and BTBD7) were spiked in the probe set as internal controls for studying viral expression.

The SARS-CoV-2 virus isolation and culturing was reported previously, which followed the CDC guidelines and good practice in laboratory health and safety requirement. Experiments were performed with approvals of W96-027B framework. Therefore, a SARS-CoV-2 virus strain (20SF014) was cultured in Vero-E6 cell, and RNA preparation performed using the QIAamp Viral RNA Mini Kit (Qiagen, Hilden, Germany). The cell culture media was collected and tested for viral RNA using the RT-PCR amplicons of partial ORF1ab with Ct value 36, and N gene regions with Ct value 33, respectively.

We divided the total RNA sample into six samples (with following slightly different experimental conditions) (Table 1). Control is RNase free water. Six samples were reverse-transcribed into cDNA, followed by the second-strand synthesis. Utilization of the synthetic double-stranded DNA, six DNA libraries were constructed through DNA fragmentation, end-repair, adaptor-ligation, and PCR amplification. Subsequently, library hybridization capture was performed by using the SARS-CoV-2 enrichment probe set. The enriched libraries were qualified with Agilent 2100 Bioanalyzer using Agilent High Sensitivity DNA Kit and equivalent double-stranded DNA libraries were pooled and transformed into a single-stranded circular DNA library through DNA-denaturation and circularisation. DNA nanoballs were generated from single-stranded circular DNA by rolling circle amplification, then qualified with Qubit 2.0 and loaded onto the flow cell and sequenced with PE100 on the MGI-2000 platform (MGI, Shenzhen, China). Detailed experimental protocol in the Chinese and English version is presented in the Supplementary Data 1.

**Table 1.** Summary statistics for six enrichment libraries from one cultured SARS-CoV-2 virus strain (20SF014)

| Sample ID | Volume<br>(ul) | PCR Circles<br>of Library<br>Amplification | Total_reads_raw | Total_reads_rmdup | Map_2019COV_Reads | nCov_Endo_Ratio | Cluster_Factor | Mean_depth |
| --- | --- | --- | --- | --- | --- | --- | --- | --- |
| 20200217A_1 | 9 | 15 | 5841044 | 5036902 | 4813734 | 0.95569 | 1.15965 | 15070.33 |
| 20200217A_2 | 9 | 15 | 5738560 | 4797881 | 4412665 | 0.91971 | 1.19606 | 13816.72 |
| 20200217A_3 | 7 | 15 | 6125219 | 5150484 | 4638866 | 0.90067 | 1.18925 | 14526.37 |
| 20200217A_4 | 7 | 15 | 8837914 | 7432367 | 6694515 | 0.90072 | 1.18911 | 20966.49 |
| 20200217A_5 | 6 | 17 | 170189070 | 150142532 | 145007261 | 0.96580 | 1.13352 | 454007.04 |
| 20200217A_6 | 7 | 17 | 224071046 | 199421414 | 192545532 | 0.96552 | 1.12361 | 602829.61 |
| 20200217A | - | - | 420802853 | 371981580 | 358112573 | 0.93469 | 1.1652 | 1121216.55 |

The cutadapt (version 2.7) and trimmonmatic (version 0.38) software was used for clipping adaptors and trimming low-quality reads. After removing adaptor, low-quality, and low-complexity reads, high-quality reads were first filtered against the human reference genome (hg 38) using Burrows-Wheeler Alignment (MEM). The remaining non-human reads were then realigned to the SARS-COV-2 reference (MN908947.3, https://www.ncbi.nlm.nih.gov/nuccore/MN908947) using bowtie2 (version 2.3.4.1) and filtered reads according to mapping quality (-q 30) by samtools (version 1.10). The variant was called by samtools and varscan (version 2.3.9, parameter: --strand-filter 0 --min-avg-qual 30 --min-reads2 15 --min-coverage 15). Finally, the sample consensus sequence was created by samtools and bcftools (version 1.9) according to the variants called above.

The results illustrated in Table 6, for library 1-6, 4,797,881-199,421,414 unique reads were obtained. Among them, 4,412,665-192,545,532 reads were mapped to SARS-CoV-2 reference sequence (MN908947.3). The fraction of SARS-CoV-2 endogenous DNA from these enrichment libraries were found to be between 90.07% and 96.58%, demonstrating that the numbers of mapped reads to SARS-CoV-2 reference sequence significantly increased, compared to metagenomic sequencing technology, following SARS-CoV-2 probe enrichment. The library complexity is evaluated by Cluster Factor, which is defined by “the number of raw reads divided by the number of reads after removing duplicates”. In our six enrichment libraries, the Cluster Factor is about 1.1. The closer to 1, the better the library construction is. Notably, when adding the PCR cycle numbers of library amplification from 15 to 17, the library quality gets better. Collectively, merging the data from six enrichment libraries, a total of 371,981,580 reads were obtained, among which 358,112,573 reads were mapped to SARS-CoV-2 reference. Using these unique SARS-CoV-2 fragments, we reconstructed six SARS-CoV-2 genomes (mean depth being 186,869× and minimum coverage 13,816×). Only the merged sequence (coverage 1,121,217×) was used for further analysis. There are five variants called from merged data, including one homozygous variant at SNP (T23569C), and four heterozygotic variants (three SNPs: C4534T, A5522T, C23525T, and one deletion: CT16779C). The phenomenon of heterozygosity had also been reported in previous studies [6,10], but we propose that this heterogeneity could be caused by the mutations that occur during viral cultivation or the infection by multi-strain of coronavirus.

We collected the variations information (gff3 files) of hig-quality samples from The Genome Variation Map (GVM) (ftp://download.big.ac.cn/GVM/Coronavirus/gff3/) (on 2020/03/22). According to the quality criteria for 2019-nCoV delivered by National Genomics Data Center (2019nCoVR, https://bigd.big.ac.cn/ncov) (Zhao, 2020), we enrolled the 597 samples with 45 SNVs at first and second levels (with MAF>0.01 and no dense variation regions, see https://bigd.big.ac.cn/ncov/variation/annotation) in the following analysis. The information of raw variations in gff3 file is recoded into binary format as an input file for Network analysis (Network version 5, www.fluxus-engineering.com) (Supplementary Table S1). Five clades could be identified and labelled, corresponding to the Full genome tree delivered by GISAID (see Figure 1). Except for three main larger clades [named: S:ORF8-L84S (defined by SNP: 28144), G: S-D614G (SNP: 23403), V:NS3-G251V(SNP: 26144)], we defined a new clade I: orf1ab-V378I (segregating at position 1397). The haplotype of the reference genome (MN908947) is in the centre clade (yellow circle), and our sample studied here is also in this clade with only one homozygous variant at position 23569 (T→C).

**Figure 1.**
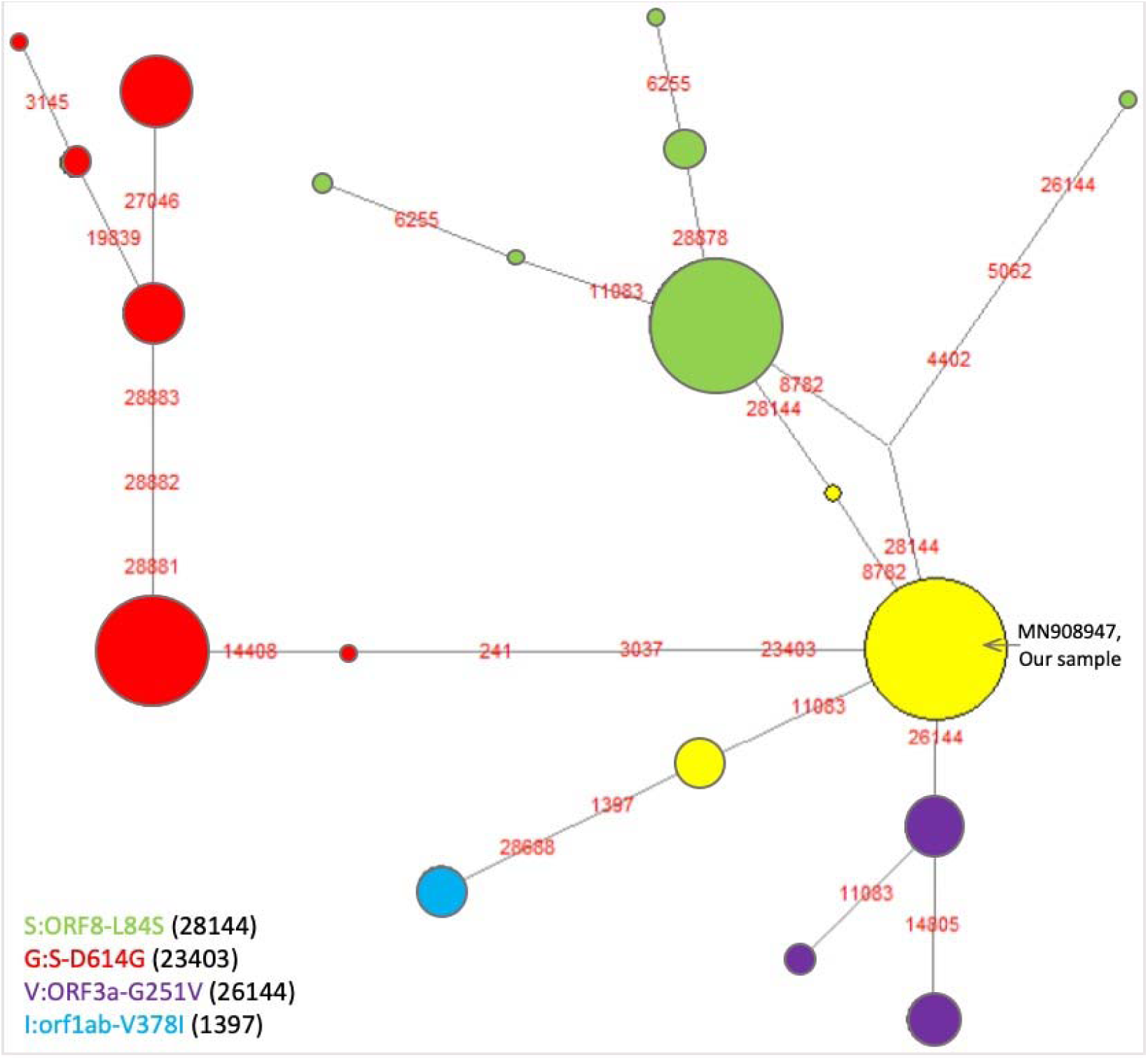
Five clades revealed by network analysis based on 597 SARS-CoV-2 genomes.

In Figure 2, we found two peaks in genome sequencing depths, one covering the 5’UTR region (MN908947.3:1-256) and another covering the N region (MN908947.3:28274-29533), which may be associated with the high expression in these two regions during replication of coronavirus [11-12]. For high sequencing depths in 5’UTR region, a reasonable explanation is that 5’UTRs before ORF1a is necessary for the discontinuous synthesis of sub genomic RNAs in the beta coronaviruses and contains the cis-acting sequences necessary for viral replication [11]. Clinically, N gene RT-PCR assay was found to be more sensitive than other genes in SARS-CoV-2 detection, which is consistent with our finding of high sequencing depths in N region. This can be explained as the structural composition of coronavirus, also the difference in expression regulation in the host cells regarding sub genomic mRNA [12-14]

**Figure 2.**
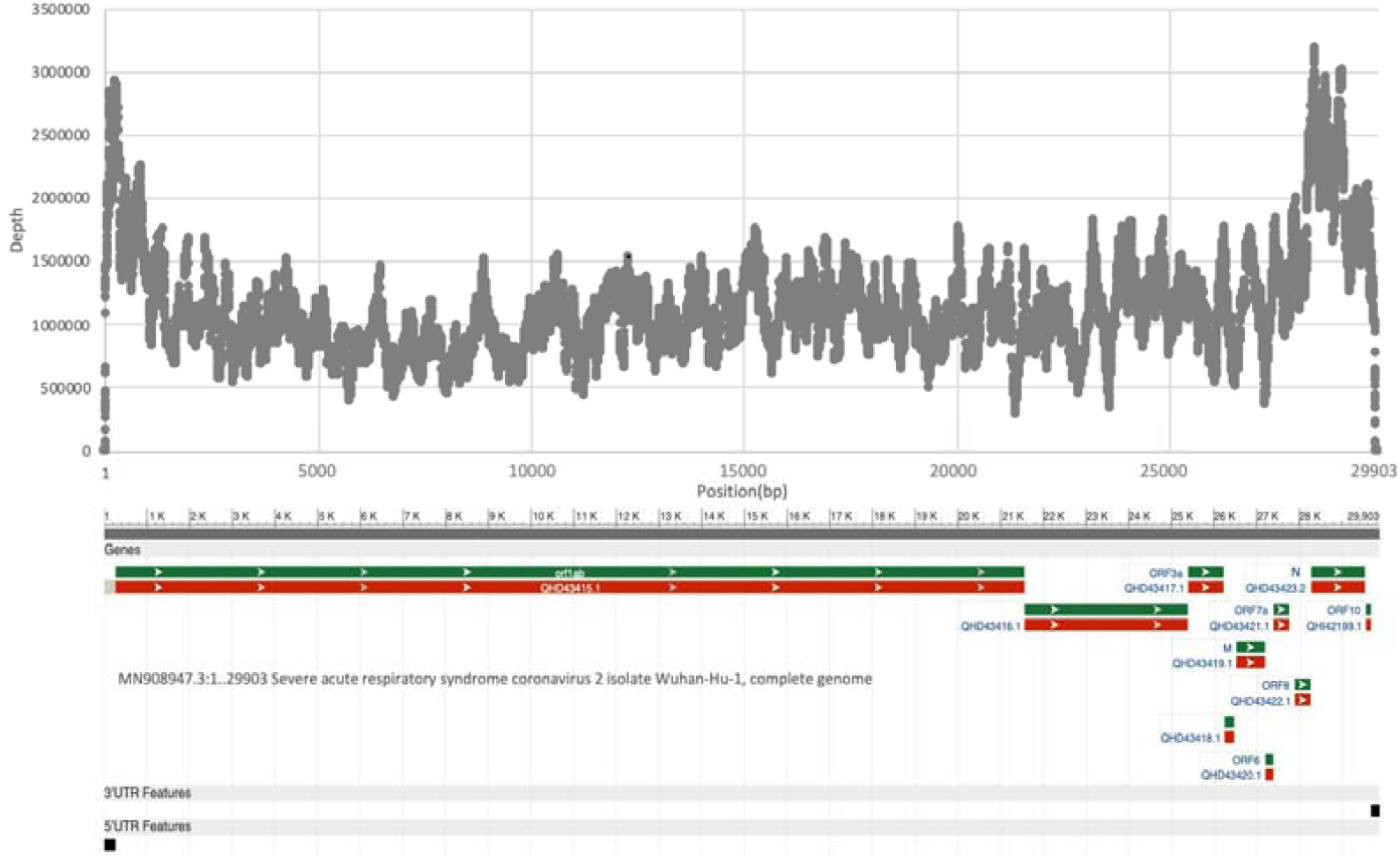
Sequencing depths corresponding to SARS-CoV-2 genome reference (MN908947.3)

In the current study, we, based on the available SARS-CoV-2 virus sequences, designed a set of SARS-CoV-2 enrichment probes. We made six enrichment libraries from one cultured SARS-CoV-2 virus strain to test the enrichment effects and sequenced them on MGI-2000 platform. Overall, the SARS-CoV-2 enrichment probe set-up described in this study showed significance in SARS-CoV-2-specific enrichment. This high specific probe rate with RNA carrier-free approach can function as a useful tool for the SARS-CoV-2 research community, especially in detecting SARS-CoV-2 RNA in low amounts and monitoring the virus evolution continuously.

## Data Availability

No external dataset

## ACKNOWLEDGEMENTS

We sincerely thank those who are on the front lines battling SARS-CoV-2 virus. We also thank the technical support provided by Guangzhou Koalson Bio-Technique Co. Ltd. Groups interested in testing this protocol can request guidance by emailing, and a limited number of our SARS-CoV-2 enrichment probe set are available on request.

## Electronic supplementary materials

**Doc S1** A protocol for in-solution SARS-CoV-2 genome capture enrichment (PDF)

**Table S1** The information of 597 high-quality SARS-CoV-2 sequences (especially 45 selected SNVs and their weight) for Network analysis in this study (XLS)

## REFERENCES

1. Chan JF, Yuan S, Kok KH et al (2020) A familial cluster of pneumonia associated with the 2019 novel coronavirus indicating person-to-person transmission: a study of a family cluster. Lancet. 395(10223):514–523.

2. Ai T, Yang Z, Hou H et al (2020) Correlation of Chest CT and RT-PCR Testing in Coronavirus Disease 2019 (COVID-19) in China: A Report of 1014 Cases. Radiology. 26:200642.

3. Wu F, Zhao S, Yu B et al (2020) A new coronavirus associated with human respiratory disease in China. Nature. 579(7798):265–269.

4. Zhou P, Yang XL, Wang XG et al (2020) A pneumonia outbreak associated with a new coronavirus of probable bat origin. Nature. 579(7798):270–273.

5. Lu R, Zhao X, Li J et al (2020) Genomic characterization and epidemiology of 2019 novel coronavirus: implications for virus origins and receptor binding. Lancet. 395(10224):565–574.

6. Tang XL, Wu CC, Li X et al (2020) On the origin and continuing evolution of SARS-CoV-2. National Science Review. waa036, https://doi.org/10.1093/nsr/nwaa036.

7. Tewhey R, Nakano M, Wang X et al (2009) Enrichment of sequencing targets from the human genome by solution hybridization. Genome Biol. 10(10):R116.

8. Briese T, Kapoor A, Mishra N et al (2015) Virome Capture Sequencing Enables Sensitive Viral Diagnosis and Comprehensive Virome Analysis. mBio. 6(5):e01491–15.

9. Zhang H, Zheng HY, Zou LR et al (2020) First Isolation and Identification of SARS-CoV-2 in Guangdong Province, China. Chinese Journal of Virology. https://doi.org/10.13242/j.cnki.bingduxuebao.003657.

10. Chen L, Liu W, Zhang Q et al (2020) RNA based mNGS approach identifies a novel human coronavirus from two individual pneumonia cases in 2019 Wuhan outbreak. Emerg Microbes Infect. 9(1):313–319.

11. Yang D, Leibowitz JL. (2015) The structure and functions of coronavirus genomic 3’ and 5’ ends. Virus Res. 206:120–33.

12. Chu DKW, Pan Y, Cheng SMS et al (2020) Molecular Diagnosis of a Novel Coronavirus (2019-nCoV) Causing an Outbreak of Pneumonia. Clin Chem. pii: hvaa029.

13. Zijie Shen, Yan Xiao, Lu Kang et al (2020) Genomic diversity of SARS-CoV-2 in Coronavirus Disease 2019 patients. Clinical Infectious Diseases, ciaa203.

14. Changtai Wang, Zhongping Liu, Zixiang Chen et al (2020) The establishment of reference sequence for SARS-CoV-2 and variation analysis. J Med Virol, 2020;1–8.

